# Caregiving Demands and Depression Symptoms among Caregivers of Individuals with Down Syndrome during the COVID-19 Pandemic

**DOI:** 10.64898/2026.05.20.26353699

**Authors:** Jennifer Nguyen, Carla Wall, Ester Jo, Lindsay Allen, Naomi Wheeler, Nicole Baumer, Allison D’Aguilar, Timothy P. York, George Capone, Colleen Jackson-Cook, Ananda B. Amstadter, Ruth C. Brown

**Affiliations:** Virginia Commonwealth University, Department of Psychiatry; Palm Beach Atlantic University, Department of Counselor Education; Virginia Commonwealth University, Department of Counseling & Special Education; University of Colorado, Department of Pediatrics; Virginia Commonwealth University, Department of Human & Molecular Genetics; Johns Hopkins University, Kennedy Krieger Institute; Virginia Commonwealth University, Departments of Pathology and Human & Molecular Genetics

**Keywords:** Down syndrome, caregiver depression, COVID-19, social determinants

## Abstract

**Background:** This study examined the association between caregiving demands and depression symptoms among caregivers of individuals with Down syndrome during the COVID-19 pandemic.

**Method:** We conducted an online survey of 200 caregivers of children and adults with Down syndrome, including demographic data, the Patient Health Questionnaire-8 (PHQ-8), and questions about lack of childcare and taking over instruction during the pandemic. A multiple linear regression analysis identified predictors of caregiver depression symptoms.

**Results:** Household income (B = −3.45, p < .001) and having to take over instruction (B = 2.24, p < .001) were significant predictors of PHQ-8 scores. Child age, caregiver gender, difficulty paying for health insurance, and lack of childcare were not significant predictors.

**Conclusions:** Lower income and instructional caregiving demands were associated with higher depression symptoms among caregivers of individuals with Down syndrome, suggesting potential targets for policy and intervention during future public health emergencies.

## Introduction

Down syndrome is a chromosomal disorder characterized by an additional chromosome 21 (Trisomy 21) that occurs in approximately 1 in 700 live births in the US (Centers for Disease Control, 2024). Down syndrome is associated with several health and developmental concerns, including higher rates in certain medical conditions, delays in language, motor, and adaptive skills, and intellectual disability (Centers for Disease Control, 2024). Although caregivers of children with developmental disabilities, including Down syndrome, report many positive aspects of parenting a child with a disability (Beighton & Wills, 2017; Hastings & Taunt, 2002; Mitchell et al., 2015), the additional responsibilities of caregiving a child with special healthcare and educational needs have been shown to negatively impact the caregiver’s mental and physical health (Dias et al., 2022, Russell et al., 2020; Scherer et al., 2019; Xia et al., 2023). These challenges have been highlighted by the U.S. Surgeon General’s advisory (2024) which reported that parents of children with disabilities face unique and heightened stressors, such as learning specialized caregiving skills, navigating complex support systems, and managing significant financial pressures.

Although there are limited studies, studies have suggested that abrupt changes in routines and services during the COVID-19 pandemic led to heightened emotional and behavioral issues among individuals with intellectual and developmental disabilities, including adults with Down syndrome (Costa et al., 2022; Villani et al., 2020) which may adversely impact caregivers. Willner et al. (2020) found that carers of children with intellectual disabilities experienced higher anxiety and depression, and lower social support compared to those without disabilities during the pandemic. Parents reported challenges such as financial instability, unmet educational and developmental needs, health concerns, and behavioral issues due to disrupted routines. Similarly, caregivers of adults with Down syndrome faced negative impacts on healthcare access, social activities, and mental health (Rubenstein et al., 2023), while shouldering increased caregiving duties due to school closures and lack of support services (Alshatti et al., 2021). Research has also documented the emotional impact of the pandemic on individuals with Down syndrome themselves, including heightened anxiety and social-related worries (Sideropoulos, Kye, et al., 2022; Sideropoulos, Sokhn, et al., 2023), and caregivers of children with learning disabilities reported using varied coping strategies to manage pandemic-related stress (Steindorsdottir et al., 2024). Together, these studies suggest that limited external resources, lack of childcare, unmet educational needs, and unavailable support services, left many caregivers vulnerable to caregiver stress, depression, and anxiety during the pandemic (Hielscher et al., 2022; U.S. Surgeon General’s Advisory, 2024).

### The Present Study

Together, existing studies suggest that the pandemic negatively affected the wellbeing of people with Down syndrome and likely their caregivers. In light of the Surgeon General’s emphasis on the unique stressors faced by these parents, more research is needed to understand the factors associated with depression among caregivers of people with Down syndrome during a public health emergency. Hence, the present study aimed to examine individual differences in depression symptoms among caregivers of people with Down syndrome in the United States during the COVID-19 pandemic, and to evaluate whether social determinants of health and pandemic-related caregiving demands, specifically lack of childcare and having to take over instruction, explained variation in those symptoms after controlling for the phase of the pandemic in which data were collected.

## Methods

### Participants

Participant data were drawn from a larger mental health surveillance study of parents and caregivers of children or adults with Down Syndrome at BLINDED FOR PEER REVIEW. Participants included 200 caregivers of children and adults with Down syndrome who completed an online survey, passed data quality checks, and completed the PHQ-8 depression screen. Inclusion criteria for the overarching study were: (a) primary caregiver of a person with Down syndrome; (b) caregiver over the age of 18; (c) able to read English or Spanish and (d) residence in the United States.

Study descriptions and recruitment flyers were distributed by Down syndrome advocacy organizations via their respective websites, social media pages, and internal listservs. Identical English and Spanish versions of the recruitment materials were used and shared in the same manner. Recruitment materials were also shared through the DS-Connect registry (Peprah et al., 2015).

### Procedures

The study was approved by the Institutional Review Board at BLINDED FOR PEER REVIEW. An online survey of caregivers of individuals with Down syndrome was distributed from October 2020 through April 2022 in REDCap (Research Electronic Data Capture; Harris et al., 2019).

### Measures

#### Demographics

Following enrollment, caregivers completed more detailed demographic questions including gender (including man, woman, non-binary, transgender, none, and prefer not to answer), race (including American Indian/Alaskan Native, Asian, Black/African American, Native Hawaiian/Other Pacific Islander, White, Other), ethnicity (Hispanic/Non-Hispanic), household income, and trouble paying for insurance.

#### Caregiving Demands

Caregiver demands were measured using two items, “Had to take over teaching child,” and “Childcare unavailable” from the Epidemic-Pandemic Impacts Inventory (EPII; Grasso, Briggs-Gowan, Carter, Goldstein, Ford, 2021). Respondents were asked to indicate for each statement whether the pandemic has impacted them or their family. The original response scale for each item was limited to Yes (me), Yes (person in home), No, and N/A. We added an additional response option of Yes (person with Down syndrome).

#### Depression

The Patient Health Questionnaire-8 (PHQ-8; Kroenke et al., 2009) is an 8-item Likert scale measure for symptoms of depression, with an additional item asking about the impact of the symptoms. Caregivers were asked to report their own depression symptoms over the past two weeks. Total scores range from 0 to 27, and scores above 10 are predictive of the presence of depressive disorders in the general population. In the current sample, Cronbach’s alpha was α = 0.87.

### Data Analysis Plan

Quality control measures were implemented to screen data for low-quality and/or falsified responses based on completion time, straight-lining answers, and endorsing multiple improbable combinations of answers (Goodrich et al., 2023). Analyses were conducted using the R Statistical language (version 4.4.1; R Core Team, 2024) on Windows 10 x64. Correlations were reported using apa.cor.table() in the `apaTables` library using two-tailed p-value cutoffs on pairwise complete observations (Stanley, 2021). To examine the association between caregiving demands, social determinants of health, and caregiver depression symptoms, we conducted an ordinary least squares linear regression using the lm() function to predict PHQ-8 score based on age of person with Down syndrome, caregiver race (White vs. non-White), household income, trouble paying for insurance, and the two EPII items “had to take over teaching child” and “childcare unavailable” as independent variables. To address the heterogeneity of the pandemic experience across the 18-month data collection period, we created a Wave variable categorizing participants by time of survey completion: Wave 1 (October-November 2020; 6.8 – 7.3 months), Wave 2 (March – July 2021; 12.3 – 15.6 months), and Wave 3 (December 2021 – March 2022; 20.9 – 25.0 months post-pandemic onset), and we included this as a covariate in the final model.

## Results

A total of 200 participants were included in the analysis following data quality screening and completion of the PHQ-8. Participant demographic information is presented in Table 1 and shows the sample majority identified as White non-Hispanic (92%), well-educated, and with higher income (54% reporting household income above $100,000). The majority of caregivers identified as female (n = 163; 82%). Mean age of the person with Down syndrome was 15.29 years (SD = 10.76). Data were collected across three waves: Wave 1 (0–10 months post-pandemic onset; n = 47, 24%), Wave 2 (10–20 months; n = 68, 34%), and Wave 3 (20–30 months; n = 85, 43%). Median PHQ-8 scores declined across waves (Wave 1: Mdn = 6; Wave 2: Mdn = 4; Wave 3: Mdn = 3; p = .006), and the proportion of caregivers who reported taking over instruction trended downward (68%, 60%, 48%; p = .070). Overall, a majority of caregivers (57%) expressed that they had to take over child instruction during the COVID-19 pandemic and 20% indicated childcare was unavailable.

**Table 1.** Means, standard deviations, and correlations.

| Variable | *M* | *SD* | 1 | 2 | 3 | 4 | 5 | 6 | 7 | 8 | 9 | 10 | 11 | 12 |
| --- | --- | --- | --- | --- | --- | --- | --- | --- | --- | --- | --- | --- | --- | --- |
| 1. Child Age | 15.20 | 10.67 |  |  |  |  |  |  |  |  |  |  |  |  |
| 2. Caregiver Gender, Male (N, %) | 40 | 19% | -.18** |  |  |  |  |  |  |  |  |  |  |  |
| 3. AIAN (N, %) | 2 | 0.9% | -.09 | .20** |  |  |  |  |  |  |  |  |  |  |
| 4. Asian (N, %) | 8 | 3.7% | -.04 | -.03 | -.02 |  |  |  |  |  |  |  |  |  |
| 5. Black (N, %) | 3 | 1.4% | .03 | -.06 | -.01 | -.02 |  |  |  |  |  |  |  |  |
| 6. NHPI (N, %) | 2 | .9% | -.04 | .08 | -.01 | -.02 | -.01 |  |  |  |  |  |  |  |
| 7. White (N, %) | 197 | 92% | -.01 | -.08 | -.33** | -.58** | -.26** | -.33** |  |  |  |  |  |  |
| 8. Other Race (N, %) | 2 | .9% | .13 | .08 | -.01 | -.02 | -.01 | -.01 | -.15* |  |  |  |  |  |
| 9. Difficulty Paying for Health Insurance | 0.20 | 0.40 | -.18** | .28** | .20** | -.04 | .04 | .07 | -.12 | -.03 |  |  |  |  |
| 10. Income (Median) | >\$100k | 53% | .02 | -.02 | -.09 | .08 | -.00 | -.09 | .10 | -.06 | -.18* | | | |
| 11. Take Over Instruction (N, %) | 122 | 57% | -.41** | -.12 | -.01 | .02 | -.06 | .08 | -.01 | -.01 | .00 | .02 |  |  |
| 12. No Childcare (N, %) | 43 | 20% | -.18* | -.09 | .19** | -.10 | .14* | .07 | -.07 | -.05 | .19** | -.18* | .15* |  |
| 13. PHQ 8 Total | 5.05 | 4.46 | -.26** | .04 | .03 | -.07 | -.09 | .10 | .07 | -.08 | .23** | -.33** | .31** | .25** |

*Note.* AIAN = American Indian or Alaskan Native, NHPI = Native Hawaiian/Pacific Islander
*M* and *SD* are used to represent mean and standard deviation, respectively. Values in square brackets indicate the 95% confidence interval for each correlation. The confidence interval is a plausible range of population correlations that could have caused the sample correlation (Cumming, 2014). \* indicates $p < .05$ . \*\* indicates $p < .01$ .

Bivariate correlations (Table 2) revealed that higher PHQ-8 Total scores were associated with younger age of person with Down syndrome (r = −.26, p < .01), lower income (r = −.33, p < .01), difficulty paying for health insurance (r = .23, p < .01), taking over instruction duties (p < .01), and lack of childcare (p < .01).

**Table 2.** Linear Multiple Regression of Social Determinants of Health, Pandemic Phase, and Caregiving.

| **Term** | **b** | **95% CI** | **p-value** |
| --- | --- | --- | --- |
| Intercept | 2.74 | [1.51, 3.96] | .072 |
| Wave (linear) | -1.00 | [-2.18, 0.18] | .096 |
| Wave (quadratic) | 0.38 | [-0.73, 1.48] | .502 |
| Child Age | -0.05 | [-0.12, 0.02] | .204 |
| White | 2.74 | [0.45, 5.02] | .019 |
| Caregiver Gender (Male) | 0.27 | [-1.36, 1.91] | .744 |
| Income (linear) | -3.45 | [-5.32, -1.58] | < .001 |
| Income (quadratic) | 0.06 | [-1.67, 1.79] | .947 |
| Income (cubic) | -1.29 | [-3.15, 0.57] | .174 |
| Income (quartic) | -0.76 | [-2.42, 0.89] | .362 |
| Difficulty Paying for Health Insurance | 1.55 | [-0.01, 3.11] | .051 |
| Had to Take Over Instruction | 2.24 | [0.93, 3.54] | < .001 |
| No Childcare | 0.99 | [-0.51, 2.50] | .194 |

*Note.* For caregiver gender, female is reference group. Wave 1 = 6.8–7.3 months post-pandemic onset (October – November 2020); Wave 2 = 12.3–15.6 months (March – July 2021); Wave 3 = 20.9–25 months (December 2021 – March 2022). For Wave, polynomial contrasts were used. CI = Confidence Interval. Adjusted $R^2$ = .26

The final model, which included the Wave covariate to account for the timing of data collection, yielded an adjusted R-squared of 0.267, indicating that the set of predictors accounted for about 27% of the variability in depression scores. The Wave covariate was not a statistically significant predictor of PHQ-8 scores (linear trend: B = −1.00, p = .096), suggesting that the timing of data collection within the pandemic did not significantly explain variance in depression scores beyond the other predictors. Household income remained the strongest predictor (B = − 3.45, p < .001), and taking over instruction during the pandemic remained a statistically significant and positive predictor (B = 2.24, p < .001) of PHQ-8 scores. White race was a significant predictor (B = 2.74, p = .019), though this finding should be interpreted cautiously given the small number of non-White participants (n = 16). Child age was no longer significant after controlling for Wave (B = −0.05, p = .204), suggesting that the original age effect may have been partially confounded with the timing of data collection. Caregiver gender, difficulty paying for insurance, and lack of childcare were not significant predictors of PHQ-8 scores in the final model.

## Discussion

The present study examined individual differences in depression symptoms among caregivers of people with Down syndrome during the COVID-19 pandemic, and evaluated whether social determinants of health and pandemic-related caregiving demands explained variation in those symptoms after controlling for the phase of the pandemic. Our findings partially supported our hypotheses: lower household income and having to take over academic instruction were associated with higher depressive symptom scores, and these associations persisted after accounting for the timing of data collection. Studies indicate that caring for younger children requires more time and assistance, which is associated with increased caregiver stress (Nam & Park, 2017), which may then decrease as caregivers adapt (Kózka & Przybyła-Basista, 2018). Notably, the effect of child age on caregiver depression, which was significant in bivariate analyses, was no longer significant after controlling for the timing of data collection. This may reflect the confounding of child age with data collection wave: school-age children predominated in Waves 1 and 2 (63–65% of those waves), whereas adults with Down syndrome predominated in Wave 3 (58%). Thus, the apparent age effect in the original model may have partially captured differences in the pandemic context rather than a direct relationship between child age and caregiver depression. Further, the lack of available resources and need to take over instruction further compounds challenges faced by caregivers (Chaturvedi et al., 2021). These results are consistent with prior work demonstrating that socioeconomic factors and social determinants of health exacerbate the mental health challenges faced by caregivers during the pandemic (Russell et al., 2020). By contrast, factors such as difficulty paying for insurance, and a lack of childcare were not significantly associated with caregiver depression, after controlling for the other predictors. The non-significant associations may be attributable to the demographics of the study sample (i.e., predominately high-income, well-educated, white non-Hispanic women) and the small number of caregivers (20%) who endorsed challenges with insurance or childcare.

Notably, the association between instructional caregiving demands and depression symptoms persisted after controlling for the phase of the pandemic in which data were collected, suggesting that these risk factors are not simply artifacts of the acute crisis period. These findings have implications for public health emergency preparedness: caregivers of children with Down syndrome who have lower incomes and who assume educational responsibilities during service disruptions may be at elevated risk for depression regardless of the specific phase of a public health emergency. Proactive identification of and support for these caregivers should be incorporated into future emergency planning for the intellectual and developmental disabilities community.

## Limitations

Results of this study should be considered within the context of several limitations. First, the study is cross-sectional with no pre-pandemic baseline measure of caregiver depression, which precludes causal inferences about the pandemic’s effect on depression and limits conclusions to associations between variables measured at a single time point. Although we controlled for the timing of data collection by including a Wave covariate, the data were collected over an 18-month period (October 2020 through April 2022) during which the pandemic context shifted substantially, from pre-vaccine lockdowns to vaccine availability and service reopening. State-level variation in pandemic policies was not captured, as geographic data beyond US residence were not collected. Second, participants were recruited through Down syndrome advocacy organizations, which likely resulted in a convenience sample biased toward more engaged and resourced families. The sample was predominantly White (92%), female (82%), well-educated, and high-income (54% reporting household income above $100,000), which substantially limits generalizability. The non-significant findings for insurance difficulty and lack of childcare may reflect restricted range in socioeconomic variables rather than true null effects. More outreach to populations with different socioeconomic backgrounds, cultural contexts, and life experiences is needed.

## Conclusion

Our findings suggest that low-income caregivers, particularly those who had to take over academic instruction due to school closures, reported higher depression symptoms. Our findings highlight the need for integrating mental health screenings for caregivers into regular healthcare visits. Healthcare professionals often overlook caregiver mental well-being and fail to monitor depression levels adequately (van den Driessen Mareeuw et al., 2020). Potential solutions could include the provision of targeted mental health services, respite care options to provide temporary relief to full-time caregivers (especially those with younger age children), and additional funding to ensure continuity of care and educational support for children with Down syndrome (Chaturvedi et al., 2021). It is also important to address the disparities that exist in access to resources and support, which involve efforts to increase the availability of and access to community resources, particularly in lower-income areas (Russell et al., 2020). Policymakers and healthcare providers are urged to recognize this need and formulate effective strategies and policies that prioritize the needs of these families.

## Data Availability

All data produced in the present study are available upon reasonable request to the authors.

## Acknowledgments

The authors would like to thank the families that participated in this study for their willingness to share their experiences during the COVID-19 pandemic and to the many students who dedicated their time to this project.

## Funding

This study was funded by the National Institute of Child Health and Human Development under grant K08 HD092610-01.

Study data were collected and managed using REDCap electronic data capture tools hosted at Virginia Commonwealth University (Harris et al., 2009) and was supported by the National Institutes of Health under Grant UM1TR004360.

## Conflicts of Interest

The authors report there are no competing interests to declare.

## approval and consent to participate

This study was approved by the Institutional Review Board at Virginia Commonwealth University, and consent to participate was provided by all parents/legal guardians of all participants.

## Author Contributions

R.B., N.W., L.A., C.J-C., A.B., G. C., and T.Y. were responsible for study design; R.B. acquired funding, performed data analysis and statistical reporting; J.N. C.W., E.J., and R.B., contributed substantially to writing the manuscript; A.D., J.N., and E.J. contributed substantially to data collection. All authors contributed substantially to the interpretation of results and edited and approved the manuscript.

## Notes

### Competing Interest Statement

The authors have declared no competing interest.

### Author Declarations

Institutional Review Board of Virginia Commonwealth University gave ethical approval for this work.

