## Supplemental Tables 1 and 2 for "Caregiving Demands and Depression Symptoms among Caregivers of Individuals with Down Syndrome during the COVID-19 Pandemic"

**Supplementary Materials**

**Table S1.** Sample Characteristics by Data Collection Wave

| Characteristic | Wave 1 (n = 47) | Wave 2 (n = 68) | Wave 3 (n = 85) | p |
| --- | --- | --- | --- | --- |
| **Age Group** |  |  |  | < .001 |
| Child (< 12) | 29 (63%) | 44 (65%) | 13 (15%) |  |
| Adolescent (12–20) | 10 (22%) | 14 (21%) | 23 (27%) |  |
| Adult (21+) | 7 (15%) | 10 (15%) | 49 (58%) |  |
| Caregiver Gender (Male) | 7 (15%) | 21 (31%) | 9 (11%) | .004 |
| Hispanic Ethnicity | 2 (4.3%) | 3 (4.4%) | 2 (2.4%) | .700 |
| White | 46 (98%) | 58 (85%) | 80 (94%) | .045 |
| Difficulty Paying for Insurance | 9 (20%) | 21 (31%) | 10 (12%) | .014 |
| Took Over Instruction | 32 (68%) | 41 (60%) | 41 (48%) | .070 |
| No Childcare | 8 (17%) | 17 (25%) | 14 (16%) | .400 |
| PHQ-8, Mdn (IQR) | 6 (3, 8) | 4 (2, 8) | 3 (1, 6) | .006 |
| **PHQ-8 Severity** |  |  |  | .015 |
| Minimal (0–4) | 17 (36%) | 37 (54%) | 55 (65%) |  |
| Mild (5–9) | 22 (47%) | 18 (26%) | 18 (21%) |  |
| Moderate (10–14) | 7 (15%) | 10 (15%) | 11 (13%) |  |
| Mod. Severe/Severe (15+) | 1 (2.1%) | 3 (4.4%) | 1 (1.2%) |  |

Note. Wave 1 = 6.8–7.3 months post-pandemic onset (October – November 2020); Wave 2 = 12.3–15.6 months (March – July 2021); Wave 3 = 20.9–25 months (December 2021 – March 2022). Percentages are column percentages. p-values from Pearson’s chi-squared test, Fisher’s exact test, or Kruskal-Wallis rank sum test as appropriate.

**Table S2.** Sample Characteristics by Age Group of Person with Down Syndrome

| Characteristic | Child (n = 86) | Adolescent (n = 47) | Adult (n = 66) | p |
| --- | --- | --- | --- | --- |
| **Wave** |  |  |  | < .001 |
| Wave 1 | 29 (34%) | 10 (21%) | 7 (11%) |  |
| Wave 2 | 44 (51%) | 14 (30%) | 10 (15%) |  |
| Wave 3 | 13 (15%) | 23 (49%) | 49 (74%) |  |
| Caregiver Gender (Male) | 24 (28%) | 7 (15%) | 6 (9.1%) | .010 |
| Caregiver Gender (Female) | 62 (72%) | 40 (85%) | 60 (91%) | .010 |
| Hispanic Ethnicity | 6 (7.0%) | 1 (2.1%) | 0 (0%) | .049 |
| White | 80 (93%) | 43 (91%) | 60 (91%) | .800 |
| Difficulty Paying for Insurance | 23 (27%) | 11 (24%) | 5 (7.6%) | .009 |
| Took Over Instruction | 63 (73%) | 35 (74%) | 15 (23%) | < .001 |
| No Childcare | 23 (27%) | 6 (13%) | 9 (14%) | .056 |
| PHQ-8, Mdn (IQR) | 7 (2, 9) | 3 (2, 6) | 2 (1, 5) | < .001 |
| **PHQ-8 Severity** |  |  |  | .003 |
| Minimal | 33 (38%) | 29 (62%) | 47 (71%) |  |
| Mild | 33 (38%) | 13 (28%) | 12 (18%) |  |
| Moderate | 17 (20%) | 4 (8.5%) | 7 (11%) |  |
| Mod. Severe/Severe | 3 (3.5%) | 1 (2.1%) | 0 (0%) |  |

Note. Child = ages 0–11; Adolescent = ages 12–20; Adult = ages 21+. One participant was missing age data. Percentages are column percentages. p-values from Pearson’s chi-squared test, Fisher’s exact test, or Kruskal-Wallis rank sum test as appropriate.
